# An exploratory analysis of the association between moral injury and mental healthcare-seeking attitudes and behaviors in returning veterans of the Iraq and Afghanistan wars

**DOI:** 10.1101/2020.08.25.20182147

**Authors:** Chad Lewis

**Author notes:** IRB Exemption Granted under DHHS Regulatory Category II, GWU Office of Human Research Study Number 031618.

## Abstract

**Objectives:** This study examines the degree to which veterans who have experienced more “moral injury” are less likely to seek the mental healthcare services they may urgently need.

**Methods:** The sampling frame for this study included American veterans of the Iraq and/or Afghanistan wars aged 25-44. Participants were recruited into the study by posting a call for participants to social media platforms such as Facebook and LinkedIn. A web link was provided to direct them to an anonymous online survey. We then collected data assessing veterans’ combat experiences, healthcare-seeking attitudes and behaviors, and various sociodemographic data. No identifying information was collected, and an Institutional Review Board exemption was granted under DHHS Regulatory Category II by the George Washington University’s Office of Human Research Study.

**Results:** The results from the Inventory of Attitudes toward Seeking Mental Health Services (IASMHS) clearly showed that moral injury was associated with more negative mental health services-seeking attitudes in all measurable areas. Those respondents scoring higher on the Moral Injury Events Scale (MIES) were presumably less likely to acknowledge their psychological problems, more likely to have fear of anticipated stigma from loved ones if they were to seek mental health treatment, and less willing and able to seek out mental health treatment when needed.

**Conclusions:** It is probable that we will not be able to reach a significant number of veterans suffering from moral injury unless we dedicate further research in the area of developing effective techniques to recruit veterans suffering from moral injury into mental health treatment programs by taking their specific symptoms (independent of PTSD) into consideration.

## Introduction

Moral injury is a psychological construct that describes extreme and unprecedented life experiences that transgress deeply held moral beliefs and expectations – including the harmful aftermath of exposure to such events (Maguen & Litz, 2015). To gain a better understanding of what a potentially “morally injurious” event could present itself as, picture a soldier patrolling down a city street in Iraq right at the moment he spots a teenage boy lying in wait to ambush his squad with a rocket-propelled grenade. As the child emerges and aims his weapon, the soldier is put in a position where he has to make a split-second judgment that carries tremendous consequences. Should he pull the trigger and ostensibly save the lives of himself and his comrades, or should he abstain from killing a child at the peril of his brothers-in-arms?

Most people would assert that he would be well within the scope of his duties to protect himself and others from an enemy combatant, but this notion is of course much easier to justify whilst safely detached from the battlefield than it is in the heart and mind of a veteran who has constantly struggled to live with him or herself many years later. This is where the psychological construct of moral injury departs from the more commonly recognized Post-Traumatic Stress Disorder (PTSD).

To sum it up in short, according to the DSM-IV criteria, PTSD’s deleterious effects on a person’s mental health are mainly rooted in recurring experiences of fear, horror, and hopelessness – specifically those which originated in the face of actual (or threatened) trauma. However, the negative effects stemming from moral injury are rooted in guilt, shame, and anger and may result in harmful states of extreme inner conflict and turmoil (Shay, 2014; Maguen & Litz, 2015). The figure shown below illustrates some areas of overlap and delineation between the two constructs.

**Figure 1.**
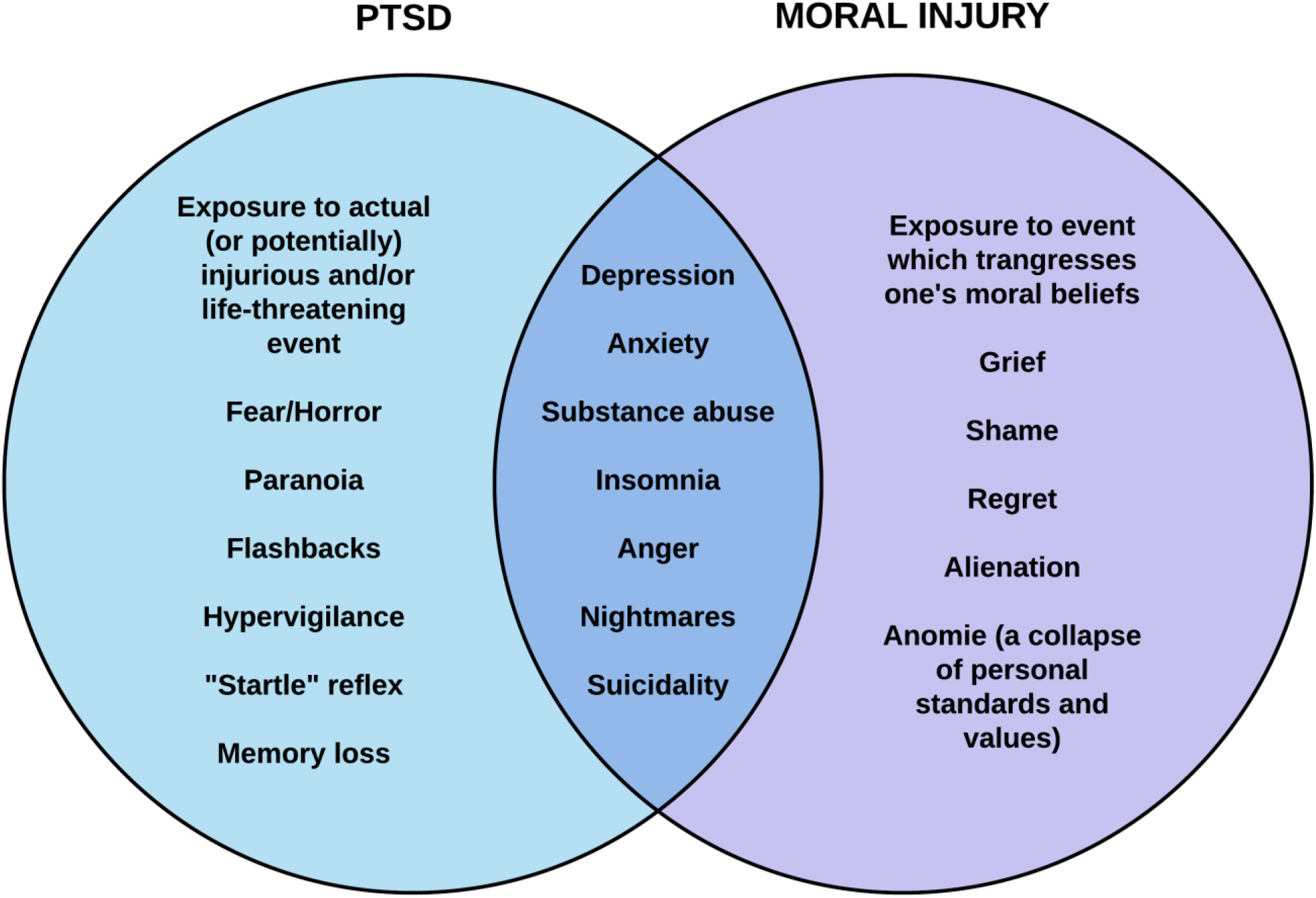
Venn diagram of PTSD and moral injury. Adapted and modified from (Wood, 2014).

While there is a paucity of epidemiological data surrounding moral injury, there is a significant amount of data on the estimated prevalence of PTSD in returning veterans. Some leaders in the field of moral injury believe that although PTSD and moral injury are fundamentally different in nature, they can both be part of the same mechanism of action. Much like a skin puncture is the primary “uncomplicated” wound, and infection is the secondary wound that provides deadlier complications over time – PTSD is usually the simpler, shorter-lived mental health outcome, whereas moral injury is an enduring impairment that can affect veterans for many years after leaving the battlefield (Shay, 2014; Severson, 2010).

For example, a study using a sample of veterans from all war eras found that 68% who screened positive for a mental health disorder did not seek any mental health treatment (Pietrzak, Goldstein, Southwick, & Grant, 2011). When applying this proportion to the RAND Center for Military Health Policy Research’s estimate of the 300,000 Iraq and Afghanistan veterans who are currently suffering from post-traumatic stress disorder or major depression, this means that approximately 204,000 of them are likely going without care (Tanelian & Jaycox, 2008). To put that number into perspective – the entire United States Marine Corps has fewer than 185,000 active duty troops (U.S. Department of Defense, 2015). Consequently, this study could carry significant implications about how to best serve the mental health needs of veterans returning from war.

After a thorough review of the published literature surrounding moral injury, it is our finding that this will be the first study to examine the relationship between moral injury and mental healthcare-seeking behaviors. Evidence pointing toward why this study is necessary includes some of the key aspects of moral injury as identified by the U.S. Department of Veterans Affairs’ National Center for PTSD. According to Maguen and Litz (2015), behavioral manifestations of moral injury may include anomie (a collapse of personal standards and values), withdrawal, self-condemnation, self-harming, and self-handicapping behaviors such as substance abuse and self-sabotage – all of which point toward a morally injured person being very unlikely to seek out help when it is needed.

## Specific Aims

The purpose of this study is to answer the following research question: “is there an association between moral injury and mental healthcare-seeking attitudes and behaviors?” The answer to this question has the potential to not only identify a need for increased awareness of moral injury as a risk factor for poor mental health outcomes, but to also inform ways in which healthcare providers, policymakers, and other stakeholders may possibly improve veterans’ health by tailoring their patient intake and surveillance systems accordingly.

Recent studies have attempted to refine the definition of moral injury, contrast moral injury against existing mental health diagnoses such as PTSD, and provide scales for measuring the extent to which returning veterans have been exposed to potentially morally injurious events. These scales – specifically the Moral Injury Events Scale (MIES) have been shown to adequately assess moral injury as it is generally defined (Nash et al., 2013; Bryan et al., 2015).

Working based on the evidence that moral injury is a valid psychological construct (Drescher et al., 2015), and that this relatively new scale is an accurate assessment tool (Nash et al., 2013; Bryan et al., 2015), this study seeks to examine the degree to which veterans who have experienced greater exposures to morally injurious events are less likely to seek the mental health services that they may urgently need.

## Methods

### Sample

The sample for the current study consisted of 229 current and former members of the American military who have served in either Iraq and/or Afghanistan. Out of the respondents, 186 (81.2%) were male and 43 (18.8%) were female. Participants ranged in age from 25 to 44 years old, with the largest age group represented being 34-37 years old (32.3%). The majority of respondents identified as White (77.2%), and the remainder of respondents identified themselves as Black (7.9%), two or more races (6.6%), “other’ (5.7%), American Indian or Alaskan Native (1.8%), or 0.9% Asian (0.9%). Nearly 17% of respondents identified as Hispanic or Latino, and one respondent failed to provide race/ethnicity information.

Just under half the sample of veterans had at least some college (27.9%) or an associate degree (19.2%), with the largest share of respondents holding bachelor’s degrees (34.1%). Out of all respondents, 28 had masters (10.9%) or professional degrees (1.3%). The majority of veterans who participated in this study were employed with 75.5% working full-time and 6.6% working part-time. Of the remaining unemployed respondents, 5.7% were students, 4.8% were not looking for work, 3.9% were disabled and unable to work, 1.7% were either keeping house or raising children full-time, and 1.7% were retired.

The majority of respondents (70%) had children under the age of 18; and most respondents were married (64%). The remaining participants were either never married (12.7%), divorced (10.1%), single but cohabitating (7.5%), separated (4.4%), or in a domestic partnership or civil union (1.3%). One respondent failed to provide relationship information and two failed to provide information on their parental status. The largest groups represented according to earnings before taxes and deductions in the 12 months prior to the study were $35K-49.9K (17.5%), $50K-74.9K (25.8%), and $75K-99.9K (22.3%). Of all respondents who elected to answer (seven did not), 21.8% earned less than $35K, and 9.6% earned greater than $100K.

Of the veterans recruited, 24.9% were currently serving in the military either through active duty (14.2%), reserve duty (7.6%), or Army/Air National Guard duty (3.1%), while 75.1% were separated form military service at the time of the survey (n=225). The mean number of deployments to Iraq or Afghanistan represented in our sample was 2.01 (n=228, SD=1.64), the mean number of years since returning from the last combat deployment was 7.64 (n=227, SD=3.94), and the mean number of years since leaving active duty (for separated respondents) was 6.66 years (n=169, SD=4.25).

When asked about their prior medical history, only 63.2% of respondents reported ever using Department of Veterans Affairs (VA) healthcare services, although 90.2% stated that they were in fact eligible for VA benefits. Fifteen respondents (6.6%) stated that they were unaware if they were eligible for VA benefits (n=228). Respondents also reported past diagnoses of PTSD (54.6%), depression (48.5%), anxiety disorder (39.3%), and traumatic brain injury (16.6%).

When asked if they ever sought mental health treatment in regards to their combat experiences (at the VA or anywhere else) such as receiving counseling or attending group therapy, 64.3% of respondents reported doing so (n=227). Twenty-seven respondents (11.8%) also reported being admitted to a hospital for psychiatric treatment at some point in the past (n=228).

### Data Collection

This study recruited a convenient sample of veterans who self-selected into the study after being provided a web link directing them to an anonymous online survey hosted by Survey Monkey. The primary locations where the link was promoted was in various veterans groups on Facebook and LinkedIn, although it is quite possible the link was shared more widely across the Internet through website posts, email, and social media messaging. The collection period lasted for 18 days between May 6 and May 24, 2016.

At the beginning of the survey, participants read a brief description of the study and provided their consent before answering a series of questions designed to gauge the nature of their combat experiences and mental healthcare-seeking behaviors through the use of various existing scales. This concluded the interaction with the participants, no follow-up was conducted.

### IRB Procedures

Before beginning the survey, participants were first asked to acknowledge that they were at least 18 years of age and capable of giving consent to participate in a study (therefore, assent and parental permission processes were not necessary). Afterward, all subjects were consented using the standard GWU Office of Human Research HRP-501 Social and Behavioral Research Consent Form. Participants provided their responses anonymously via the Survey Monkey website and were assured of their protection. Contact information for the GWU Office of Human Research and the study’s Principal Investigator, Dr. Kathleen Roche, MSW, PhD was also provided for any participants having questions about their rights as human subjects or further information on the study.

No physical forms or specimens were used. No identifying information was collected, and care was taken when designing the survey to ensure that deductive disclosure would not be possible. All participant data is currently being stored either online within Survey Monkey’s secure servers, or on password-protected desktop computers secured within research team members’ offices or private residences. Potential risks to participants in this study included any negative mental health effects that could be brought about by recalling traumatic events from their combat experiences and/or discomfort associated with disclosing their personal information.

Ultimately, this study was granted an IRB exemption under DHHS Regulatory Category II by the GWU Office of Human Research (Study #031618).

### Measures

A copy of all instruments that were used to collect the primary data reported by this study are listed in the Appendix.

#### Inventory of Attitudes toward Seeking Mental Health Services (IASMHS)

The IASMHS is a 24-item self-reported scale that was adapted and extended from Fischer and Turner’s (1970) Attitudes Toward Seeking Professional Psychological Help Scale (ATSPPHS) by researchers at the Queen’s University in Ontario, Canada in the early 2000s. The IASMHS is designed to measure respondents’ attitudes towards seeking professional psychological help, and studies have found it to be a theoretically and psychometrically superior scale to the ATSPPHS that is able to distinguish between respondents who had and had not used mental health services in the past, and those who were likely or unlikely to use mental health services in the future. It is also able to be broken down further into three subscales measuring “psychological openness,” “indifference to stigma,” and “help-seeking propensity” (Mackenzie, Knox, Gekoski, & Macaulay, 2004).

The IASMHS’ eight-item psychological openness subscale measures the extent to which respondents are willing to acknowledge having psychological problems and the possibility of seeking help for them. Scores can range from zero to 32, and a higher score indicates an individual having more likelihood of acknowledging their psychological problems and the possibility of seeking help for them (Mackenzie, et al., 2004).

The IASMHS’ eight-item indifference to stigma subscale measures the extent to which respondents place concerns upon the perceptions of others who are important to them if they were to find out the respondent was seeking mental health treatment. Scores can range from zero to 32, and a higher score indicates an individual having more likelihood of assuming their loved ones would have a negative perception of them seeking mental health treatment (Mackenzie, et al., 2004).

The IASMHS’ eight-item help-seeking propensity subscale measures the extent to which respondents believe they are willing and able to seek out mental health treatment when needed. Scores can range from zero to 32, and a higher score indicates an individual having more willingness to seek out mental health treatment when needed (Mackenzie, et al., 2004).

#### Moral Injury Events Scale (MIES)

The MIES is a nine-item self-reported scale used to measure exposure to military-connected events that violate a person’s deeply held moral beliefs and values. As an assessment tool, the MIES is designed to evaluate the existence and perceived intensity of these potentially morally-injurious combat experiences, which can help inform mental health providers with a better understanding of the psychological, biological, social, and spiritual impacts of moral injury. Scores can range from nine to 54, and a higher score indicates an individual having a greater amount of exposure to likely morally-injurious events (Nash, et al., 2013).

#### PTSD Checklist for DSM-IV – Military Version (PCL-M)

The PCL-M is 17-item self-reported scale used to measure PTSD symptoms (as outlined by the DSM-IV) by asking about symptoms in response to stressful military experiences. The PCL-M is primarily used to screen or diagnose military members and veterans for PTSD. The standard PCL (which also has two other versions, the PCL-Civilian, and PCL-Specific) was recently revised to align with the newly released DSM-5 guidelines (PCL-5). However, there is not a DSM-5 version of the PCL-M that has been developed yet. Therefore, this study employs the older, DSM-IV version of the PTSD Checklist.

Scores can range from 17 to 85, and a higher score indicates greater likelihood of PTSD diagnosis. The PCL can be scored in three various ways, but for the purposes of this study we have chosen to use the “total symptom severity score,” in which scores are summed and then compared against suggested cut-points based upon the estimated prevalence of PTSD in a specific target population (National Center for PTSD, 2014).

#### Combat Exposure Scale (CES)

The CES is a seven-item self-reported scale that was developed in the late 1980s by clinical psychologists who were experienced in the diagnosis and treatment of combat-related PTSD. The CES’ purpose is to measure self-reported wartime stressors experienced by combatants. Items on the scale are weighted according to the implied severity of the experience. For example, “being surrounded by the enemy” is weighted more heavily than “being under enemy fire,” and so on.

The scale employs varying Likert-type items ranging from one to five with differing measures for each item such as number of times, length of time, or percentage of time. The range of possible values is zero to 41, with scores ranging from zero to eight representing “light” exposure, 9-16 representing “light-moderate” exposure, 17-24 representing “moderate” exposure, 25-32 representing “moderate-heavy” exposure, and 33-41 representing “heavy” exposure (Keane et al., 1989).

## Analysis Plan

After using descriptive statistics on all variables, Cronbach’s alpha on all scales to measure internal consistency, and Shapiro-Wilk tests of normality on all scales to gain a solid understanding of the data we collected, the primary method of data analysis employed during this study in order to answer our research question is Spearman’s rank-order correlation. We chose to employ Spearman rank-order correlation instead of the more standard Pearson product-moment correlation due to the presence of non-parametrically distributed data found across all of our scales, which can be seen below in Table 1.

**Table 1.** Tests of Normality.

*Tests of Normality*
|  | Kolmogorov-Smirnov<sup>a</sup> |  |  | Shapiro-Wilk |  |  |
| --- | --- | --- | --- | --- | --- | --- |
|  | Statistic | df | Sig. | Statistic | df | Sig. |
| IASMHS Score | .070 | 201 | .018 | .980 | 201 | .007 |
| Psychological Openness Subscale Score | .099 | 206 | .000 | .976 | 206 | .001 |
| Help-Seeking Propensity subscale score | .077 | 205 | .004 | .981 | 205 | .008 |
| Indifference to Stigma Subscale Score | .106 | 202 | .000 | .959 | 202 | .000 |
| MIES Score | .069 | 198 | .022 | .972 | 198 | .001 |
| PCL-M Score | .059 | 192 | .200* | .975 | 192 | .002 |
| CES Score | .073 | 188 | .015 | .971 | 188 | .001 |

\*. This is a lower bound of the true significance.
a. Lilliefors Significance Correction

Spearman’s rho allowed us to quantify the strength of an association between ranked items pertaining to respondents’ clinical history, combat exposure, likelihood of moral injury, likelihood of PTSD, and mental healthcare-seeking attitudes.

## Results

**Table 2.** Descriptives for IASMHS (and corresponding subscales), MIES, PCL-M, and CES.

|  | N | Minimum | Maximum | Mean | Std. Deviation |
| --- | --- | --- | --- | --- | --- |
| IASMHS Score | 201 | 17 | 88 | 51.76 | 17.122 |
| Psychological Openness<br>Subscale Score | 206 | 1 | 31 | 17.49 | 6.423 |
| Indifference to Stigma<br>Subscale Score | 202 | 0 | 32 | 16.38 | 8.384 |
| Help-Seeking Propensity<br>subscale score | 205 | 4 | 28 | 17.84 | 5.515 |
| MIES Score | 198 | 9 | 54 | 28.89 | 11.458 |
| PCL-M Score | 192 | 17 | 85 | 48.77 | 17.581 |
| CES Score | 188 | 0 | 40 | 17.85 | 9.708 |
| Valid N (listwise) | 182 |  |  |  |  |

### IASMHS

Out of 201 valid responses, the mean IASMHS score was 51.76 (SD, 6.42), and scores ranged from 17 to 88. Unfortunately, due to a typographical error that wasn’t identified until the data collection process was already underway, only 103 valid responses were recorded for subscale item 22 – “I would willingly confide intimate matters to an appropriate person if I thought it might help me or a member of my family.”

This error would have required the first 126 survey responses to be excluded in order to keep the scale intact, so item 22 was dropped in order to maximize our sample size for the scale overall. This reduced the maximum possible scale score from 96 to 92. The scale also showed very strong internal reliability with a Cronbach’s alpha score of .895. When including item 22 (n=99), the Cronbach’s alpha score slightly increased to .896.

Out of 206 valid responses, the mean psychological openness score was 17.49 (SD, 6.42), and scores ranged from one to 31. The subscale also showed very strong internal reliability with a Cronbach’s alpha score of .752.

Out of 202 valid responses, the mean indifference to stigma score was 16.38 (SD, 8.38), and scores ranged from zero to 32. The subscale also showed good internal reliability with a Cronbach’s alpha score of .865.

Out of 202 valid responses, the mean help-seeking propensity score was 17.84 (SD, 5.52), and scores ranged from four to 28. The subscale also showed good internal reliability with a Cronbach’s alpha score of .752. As item 22 also belonged to this subscale, once it was dropped this subscale consisted of seven items instead of eight like the other two subscales for psychological openness and indifference to stigma. When including item 22 (n=103), the Cronbach’s alpha score did slightly increase to .755.

### MIES

Out of 198 valid responses, the mean MIES score was 28.89 (SD, 11.46), and scores ranged from nine to 54. The scale also showed very strong internal reliability with a Cronbach’s alpha score of .873.

### PCL-M

Out of 192 valid responses, the mean PCL-M score was 48.77 (SD, 17.58), and scores ranged from 17 to 85. The scale also showed extremely strong internal reliability with a Cronbach’s alpha score of .953. As stated earlier, the “total symptom severity score” is used to compare against suggested cut-points based upon the estimated prevalence of PTSD in a specific target population. Cut-point scores provide three distinct ranges in which clinicians can assess patients’ likelihood for PTSD. If the goal of the assessment is to screen, then the lower end of the range is recommended as a good cut-point in order to maximize the sensitivity of the tool. If diagnosis is the goal, then the recommended cut-point is the higher end of the range in order to minimize the likelihood of false positives identified by the tool (National Center for PTSD, 2014).

For a setting that would include general population samples, the estimated prevalence of PTSD is 15% or below, therefore the suggested cut-point scores range from 30 to 35. In settings such as those that would be found at a VA primary care clinic, a more moderate prevalence level of PTSD is assumed (16-39%), and cut-point scores range from 36-44. In a setting such as specialty mental health clinics, the highest estimated prevalence of PTSD is assumed (40% or above), and cut-point scores range from 45 to 50 (National Center for PTSD, 2014).

When scoring the PCL-M responses collected in this study using moderate PTSD prevalence assumptions, 75% of respondents met the screening criteria and 62.5% met the diagnosis criteria. When scoring using the highest PTSD prevalence assumptions, 60.4% of respondents met the screening criteria and 46.9% met the diagnosis criteria.

### CES

Out of 188 valid responses, the mean CES score was 17.85 (SD, 9.71), and scores ranged from zero to 40. The scale also showed very strong internal reliability with a Cronbach’s alpha score of .841. As explained earlier, the CES categorizes respondents’ combat exposure into five categories ranging from “light” to “heavy” (Keane et al., 1989).

When scoring the CES responses collected in this study, 14.9% of respondents were found to have experience “light” combat exposure, 31.9% experienced “light-moderate” combat exposure, 29.8% experienced “moderate” combat exposure, 13.3% experienced “moderate” combat exposure, and 10.1% experienced “heavy” combat exposure.

### Correlations

Using the correlation coefficient interpretation guidelines proposed by Evans (1996) to measure the strength of association between our scales of interest (MIES, PCL-M, and CES) and respondents’ healthcare-seeking behaviors (as interpreted from selected portions of their clinical history provided through the survey), we found many statistically significant correlations as are shown below in Table 3.

**Table 3.** Correlations between clinical history and selected scales (MIES, PCL-M, & CES)

|  |  |  | Used VA<br>healthcare<br>services | Used<br>mental<br>health<br>services | Admitted<br>for<br>psychiatric<br>treatment | Have<br>considered<br>suicide | Have<br>attempted<br>suicide | MIES<br>Score | PCL-M<br>Score | CES<br>Score |
| --- | --- | --- | --- | --- | --- | --- | --- | --- | --- | --- |
| Spearman's<br>rho | Used VA<br>healthcare<br>services | $\rho$ | 1.000 | | | | | | | |
|  |  | Sig. | . |  |  |  |  |  |  |  |
|  |  | N | 228 |  |  |  |  |  |  |  |
| | Used<br>mental<br>health<br>services | $\rho$ | .305** | 1.000 | | | | | | |
|  |  | Sig | .000 | . |  |  |  |  |  |  |
|  |  | N | 227 | 227 |  |  |  |  |  |  |
| | Admitted<br>for<br>psychiatric<br>treatment | $\rho$ | .111 | .188** | 1.000 | | | | | |
|  |  | Sig | .094 | .004 | . |  |  |  |  |  |
|  |  | N | 228 | 227 | 228 |  |  |  |  |  |
| | Have<br>considered<br>suicide | $\rho$ | .026 | .169* | .227** | 1.000 | | | | |
|  |  | Sig | .697 | .011 | .001 | . |  |  |  |  |
|  |  | N | 228 | 227 | 228 | 228 |  |  |  |  |
| | Have<br>attempted<br>suicide | $\rho$ | .106 | .130 | .292** | .359** | 1.000 | | | |
|  |  | Sig | .111 | .052 | .000 | .000 | . |  |  |  |
|  |  | N | 227 | 226 | 227 | 227 | 227 |  |  |  |
| | MIES<br>Score | $\rho$ | .123 | .305** | .192** | .382** | .361** | 1.000 | | |
|  |  | Sig | .085 | .000 | .007 | .000 | .000 | . |  |  |

|  | N | 198 | 197 | 198 | 198 | 197 | 198 |  |  |
| --- | --- | --- | --- | --- | --- | --- | --- | --- | --- |
| PCL-M Score | $\rho$ | .196** | .431** | .164* | .357** | .260** | .461** | 1.000 | |
|  | Sig | .006 | .000 | .023 | .000 | .000 | .000 | . |  |
|  | N | 192 | 191 | 192 | 192 | 191 | 191 | 192 |  |
| CES Score | $\rho$ | .223** | .284** | .098 | .105 | -.077 | .121 | .368** | 1.000 |
|  | Sig | .002 | .000 | .182 | .153 | .294 | .099 | .000 | . |
|  | N | 188 | 187 | 188 | 188 | 187 | 186 | 187 | 188 |

\*\* . Correlation is significant at the 0.01 level (2-tailed).
\* . Correlation is significant at the 0.05 level (2-tailed).

When looking at respondents’ past VA healthcare services usage, a positive correlation was found with PCL-M score (ρ=.196, p<.01), a positive correlation was also found with CES score (ρ=.223, p<.01), and no significant correlation was found with MIES score.

When looking at respondents’ history of mental health treatment in regards to their combat experiences (at the VA or anywhere else), such as receiving counseling or attending group therapy, a positive correlation was found with MIES score (ρ=.305, p<.001), a moderate positive correlation was found with PCL-M score (ρ=.431, p<.001), and a weak positive correlation was found with CES score (ρ=.284, p<.001).

When looking at respondents’ history of being admitted to a hospital for psychiatric treatment, a very weak positive correlation was found with MIES score (ρ=.192, p<.01), a very weak positive correlation was found with PCL-M score (ρ=.164, p<.05), and no significant correlation was found with CES score.

When looking at respondents’ history of considering suicide, a weak positive correlation was found with MIES score (ρ=.382, p<.001), a weak positive correlation was found with PCL-M score (ρ=.357, p<.001), and no significant correlation was found with CES score. Also, when looking at respondents’ history of attempting suicide, a weak positive correlation was found with MIES score (ρ=.361, p<.001), a weak positive correlation was found with PCL-M score (ρ=.260, p<.001), and again, no significant correlation was found with CES score.

When looking at the strength of association between our scales of interest (MIES, PCL-M, and CES) and respondents’ healthcare-seeking attitudes (as measured by the IASMHS and its three subscales), we also found many statistically significant correlations as are shown below in Table 4. When looking at respondents’ overall IASMHS scores, a weak negative correlation was found with MIES score (ρ=-.323, p<.001), a moderate negative correlation was found with PCL-M score (ρ=-.433, p<.001), and no significant correlation was found with CES score.

**Table 4.**
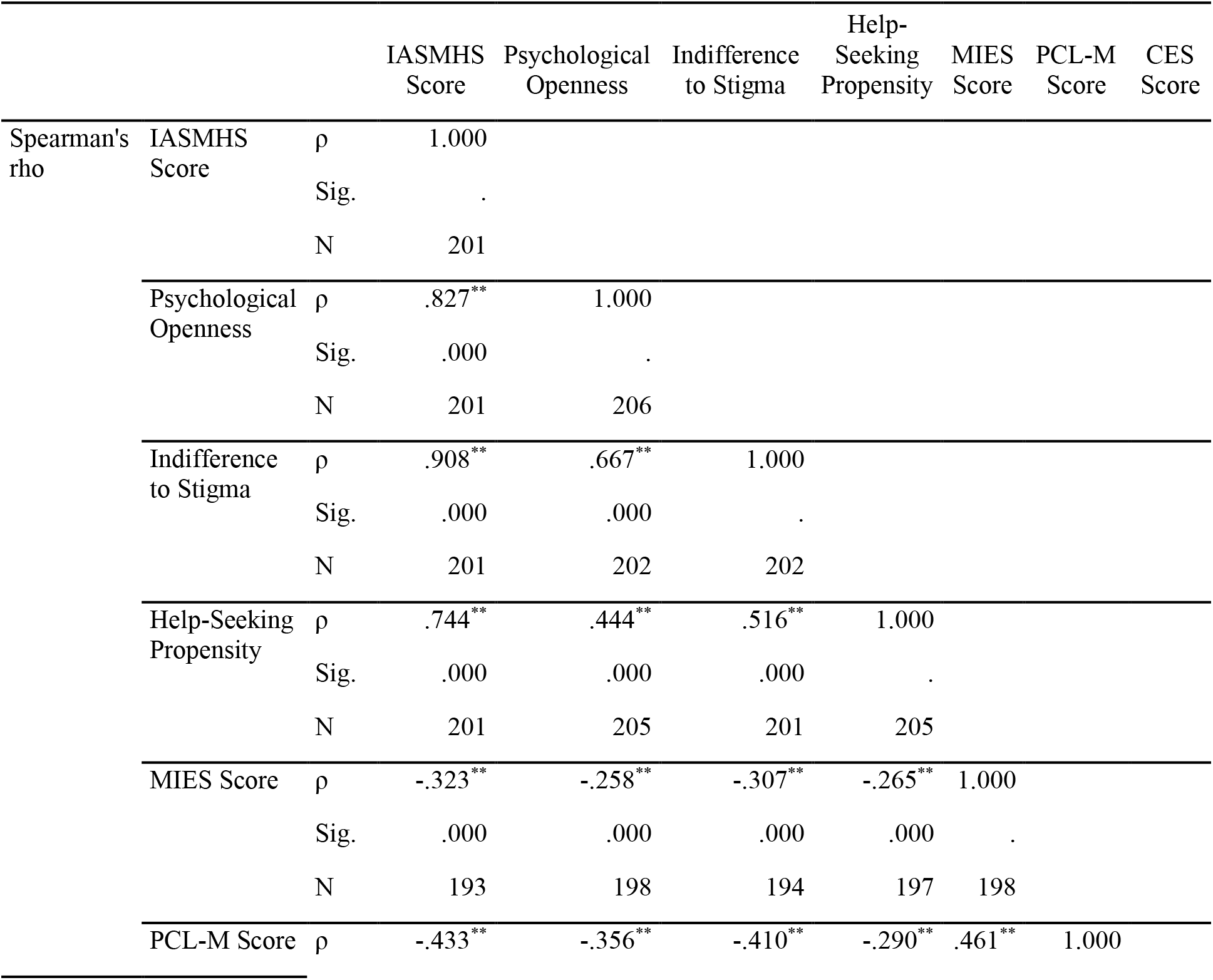
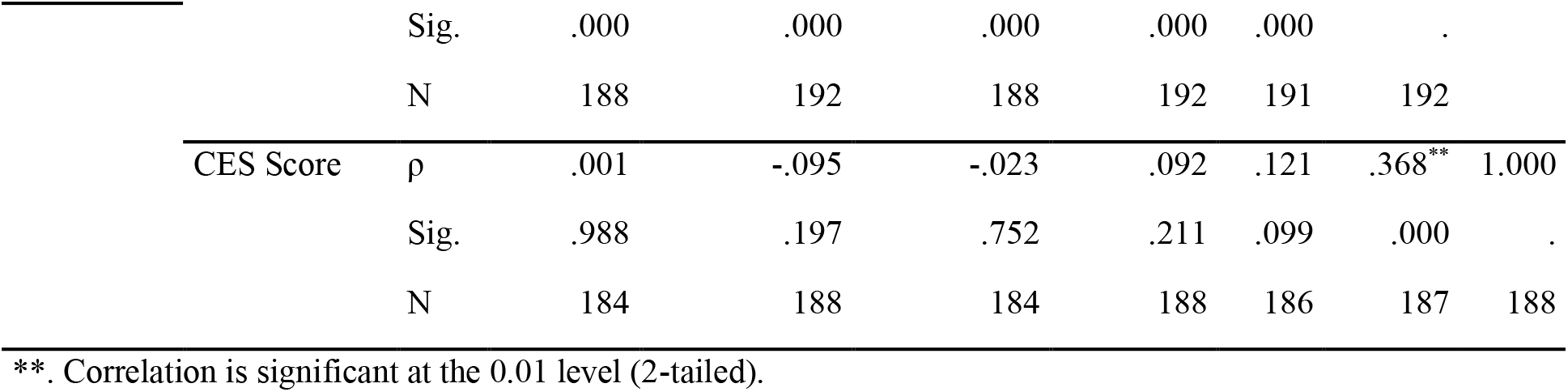
Correlations between the IASMHS (w/subscales) and selected scales (MIES, PCL-M, & CES)

Also, when looking at respondents’ psychological openness subscale scores, a weak negative correlation was found with MIES score (ρ=-.258, p<.001), a weak negative correlation was found with PCL-M score (ρ=-.356, p<.001), and no significant correlation was found with CES score. When looking at respondents’ indifference to stigma subscale scores, a weak negative correlation was found with MIES score (ρ=-.307, p<.001), a moderate negative correlation was found with PCL-M score (ρ=-.410, p<.001), and no significant correlation was found with CES score. Finally, when looking at respondents’ help-seeking propensity subscale scores, a weak negative correlation was found with MIES score (ρ=-.265, p<. 001), a weak negative correlation was found with PCL-M score (ρ=-.290, p<.001), and again, no significant correlation was found with CES score.

## Discussion

When it comes to our primary research question, “is there an association between moral injury and mental healthcare-seeking behaviors?” The answer we have found is clearly, yes. In every way that we chose to measure attitudes and behaviors pertaining to help-seeking, those scoring higher for moral injury either showed associations with outcome variables as we predicted they would, or had worse outcomes than those scoring higher for PTSD . For example, even though we found that MIES score and PCL-M score were both positively associated with whether respondents’ had used VA healthcare services (ρ=.123, p=.085 versus ρ=.196, p<.01) or mental health services (ρ=.305, p<.001 versus ρ=.431, p<.001) in the past (which we did not anticipate), those scoring higher on the MIES were less strongly associated for both measures, and therefore did not seek help at the same rate as those scoring higher for PTSD.

Also, when looking at the other clinical history outcome variables we measured, those scoring higher on the MIES had stronger positive associations with undesirable health outcomes than those scoring higher on the PCL-M including suicidal ideation (ρ=.382, p<.001 versus ρ=.357, p<.001), attempted suicide (ρ=.361, p<.001 versus ρ=.260, p<.001), and past psychiatric admittances to a hospital (ρ=.192, p<.01 versus ρ=.164, p<.05).

Surprisingly, PCL-M scores were more strongly negatively associated with all IASMHS mental healthcare-seeking attitudes measures than were MIES scores, although both had weak to moderate negative associations both overall and across all three subscales (psychological openness, indifference to stigma, and help-seeking propensity) that were significant at the .001 level. This evidence even further supports the idea that those scoring higher for moral injury would be less likely to seek mental healthcare services.

Although we were able to achieve our modest goal of recruiting at least 200 participants into this study, since it was conducted on a very limited budget and schedule, there were quite a few limitations faced when it came to recruiting a large number of participants into the study. Not only did the lack of being able to provide a financial incentive to participation most likely make it harder to recruit participants into the study, but it is also likely that the nature of the pages the survey link was shared on (specifically within the social media platforms Facebook and LinkedIn) also contributed to our relatively small sample size.

While the survey link remained active for a period of 18 days, over 93% of survey responses were received in the first seven days. We believe that this happened for one of two reasons, either the link became buried under too many other posts on the same page which prevented our recruitment post from being viewed easily by visitors to the page, or the presence of the link simply achieved its goal by becoming fully saturated and reaching everyone who was willing to self-select into the survey.

We believe that the first assumption is more likely due to the sheer number of people belonging to the groups in which the link was shared and the disproportionately low number of views the posts received. For example, the recruitment post created on LinkedIn was shared to a number of veteran- and mental health-specific groups comprised of over 340,000 (not necessarily unique) individuals, and only 1,038 views were recorded for the post. Given this experience, we believe that social media-based recruitment links require more active monitoring and refreshing of posts in order to keep the link at the “top” of people’s news feeds. It is quite possible that given the speed in which we received our first two hundred respondents that we could have doubled or even tripled the final number of survey responses received had the recruitment posts been more carefully monitored.

According to the U.S. Department of Defense’s most recent demographics report, we believe that we were mainly successful in recruiting a representative sample of participants when looking at demographic variables such as gender and race when compared to active duty personnel. Women comprise approximately 15% of active duty personnel, and our sample was almost 19% female. Slightly more than 30% of active duty personnel identify with a racial minority group and our sample was somewhat less diverse at 23%. However, where 12% of active duty personnel identify as Hispanic, nearly 17% of our respondents identified as Hispanic or Latino. As far as education and earnings are concerned, our sample was at a much higher level in both areas, however we believe that this can be expected with older, separated personnel who have been out of the military for a few years, developed their careers, and have VA educational benefits available to help them pay for college.

One of the primary areas in which our sample did not seem to be representative of the larger veteran population was their prior clinical history. Respondents reported past diagnoses of post-traumatic stress disorder (54.6%), depression (48.5%), anxiety disorder (39.3%), and traumatic brain injury (16.6%) at much higher rates than what is currently accepted as the nationwide prevalence rates. According to Gradus (2016), the prevalence of PTSD found in participants of the Gulf War Study (n=1,938) was 13.8% using the PCL as we did in our study.

When scoring our participants responses using the highest PTSD prevalence assumptions and the higher cut-off score required to meet the diagnosis criteria, our sample of veterans still had a apparent prevalence of PTSD nearly 3.4 times the amount that the veterans of the Gulf War Study had at 46.9%. We believe that due to this, our study has a significant amount of selection bias that must be taken into consideration when attempting to generalize the finding of or study. For future studies, it would improve the quality of our exploration if we were to have a sample more representative of the larger veteran population in consideration to mental health history.

When looking at the correlations between all scales (IASMHS w/subscales, MIES, PCL-M, and CES) and various demographic factors such as age, gender, education, and earnings, there were extremely few statistically significant associations found. On top of this, none of the significant correlations exceeded the “very weak” strength of association guidelines as set out by Evans (1996), therefore they were not explored further through multivariate regression analysis.

## Conclusions

Veteran suicide is a hot topic in America right now. Social media is currently ablaze with infographics and videos promoting awareness of the estimated 22 veterans who are committing suicide in the U.S. every day. According to Christenson (2016), an even more recent reminder of the devastating toll that suicide has taken on our veteran population came out of the Pentagon as the number of military suicides since 2003 (4,839) exceeded the number of troops that were killed in Iraq during the same time period (4,496).

This is especially pertinent when trying to understand why moral injury in returning veterans is such a crucial issue. According to Hendin and Hass (1991), when studying PTSD manifestation in Vietnam veterans, they found that combat-related guilt was the most significant predictor of suicide attempts and suicidal ideations. Since guilt is essentially the primary facet of the psychological construct of moral injury, and yet not a condition that even needs to be present for a diagnosis of PTSD, it stands to reason that further research is necessary to strengthen the case for moral injury as a valid construct that needs widespread acceptance and support within the mental health community. This is especially important when considering the findings of this study given that those scoring higher on the MIES had stronger positive associations with both suicidal ideation and attempted suicide than those scoring higher on the PCL-M.

While this study has found that moral injury can be positively associated with whether a person has sought treatment in the past, this is very likely to have been confounded by selection bias due to the fact that our sample of respondents had a much higher prevalence of PTSD than that estimated in the veteran population. Even so, the results from the IASMHS clearly showed that moral injury was associated with more negative mental health services-seeking attitudes in all measurable areas. Those respondents scoring higher on the MIES were presumably less likely to acknowledge their psychological problems, more likely to have fear of anticipated stigma from loved ones if they were to seek mental health treatment, and less willing and able to seek out mental health treatment when needed.

For these reasons, it is probable that we will not be able to reach a significant number of veterans suffering from moral injury unless we dedicate further research in the area of developing effective techniques to recruit veterans suffering from moral injury into mental health treatment programs by taking their specific symptoms (independent of PTSD) into consideration.

## Data Availability

All participant data is currently being stored either online within Survey Monkey's secure servers, or on password-protected desktop computers secured within research team members' offices or private residences.

## Appendix

### Inventory of Attitudes toward Seeking Mental Health Services (IASMHS)

The term *professional* refers to individuals who have been trained to deal with mental health problems (e.g., psychologists, psychiatrists, social workers, and family physicians). The term *psychological problems* refers to reasons one might visit a professional. Similar terms include *mental health concerns, emotional problems, mental troubles*, and *personal difficulties*.

For each item, indicate whether you *disagree* (0), *somewhat disagree* (1), *are undecided* (2), *somewhat agree* (3), *or agree* (4):

1. There are certain problems which should not be discussed outside of one’s immediate family.
2. I would have a very good idea of what to do and who to talk to if I decided to seek professional help for psychological problems.
3. I would not want my significant other (spouse, partner, etc.) to know if I were suffering from psychological problems.
4. Keeping one’s mind on a job is a good solution for avoiding personal worries and concerns.
5. If good friends asked my advice about a psychological problem, I might recommend that they see a professional.
6. Having been mentally ill carries with it a burden of shame.
7. It is probably best not to know everything about oneself.
8. If I were experiencing a serious psychological problem at this point in my life, I would be confident that I could find relief in psychotherapy.
9. People should work out their own problems; getting professional help should be a last resort.
10. If I were to experience psychological problems, I could get professional help if I wanted to.
11. Important people in my life would think less of me if they were to find out that I was experiencing psychological problems.
12. Psychological problems, like many things, tend to work out by themselves.
13. It would be relatively easy for me to find the time to see a professional for psychological problems.
14. There are experiences in my life I would not discuss with anyone.
15. I would want to get professional help if I were worried or upset for a long period of time.
16. I would be uncomfortable seeking professional help for psychological problems because people in my social or business circles might find out about it.
17. Having been diagnosed with a mental disorder is a blot on a person’s life.
18. There is something admirable in the attitude of people who are willing to cope with their conflicts and fears without resorting to professional help.
19. If I believed I was having a mental breakdown, my first inclination would be to get professional attention.
20. I would feel uneasy going to a professional because of what some people would think.
21. People with strong characters can get over psychological problems by themselves and would have little need for professional help.
22. I would willingly confide intimate matters to an appropriate person if I thought it might help me or a member of my family.
23. Had I received treatment for psychological problems, I would not feel that it ought to be “covered up.”
24. I would be embarrassed if my neighbor saw me going into the office of a professional who deals with psychological problems.

### Moral Injury Event Scale (MIES)

Considering your active duty service during warzone deployment, indicate for each item, indicate whether you *strongly agree* (1), *moderately agree* (2), *slightly agree* (3), *slightly disagree* (4), *moderately disagree* (5), or *strongly disagree* (6):

1. I saw things that were morally wrong
2. I am troubled by having witnessed others’ immoral acts
3. I acted in ways that violated my own moral code or values
4. I am troubled by having acted in ways that violated my own morals or values
5. I violated my own morals by failing to do something that I felt I should have done
6. I am troubled because I violated my morals by failing to do something that I felt I should have done
7. I feel betrayed by leaders who I once trusted
8. I feel betrayed by fellow service members who I once trusted
9. I feel betrayed by others outside the U.S. military who I once trusted
10. I trust my leaders and fellow service members to always live up to their core values
11. I trust myself to always live up to my own moral code

### PTSD Checklist for DSM-IV (PCL-M)

Below is a list of problems and complaints that veterans sometimes have in response to stressful military experiences. Please read each one carefully, then circle one of the numbers to the right to indicate how much you have been bothered by that problem in the past month. (1 = Not at all, 2 = A little bit, 3 = Moderately, 4 = Quite a bit, 5 = Extremely)

1. Repeated, disturbing *memories, thoughts*, or *images* of a stressful military experience?
2. Repeated, disturbing *dreams* of a stressful military experience?
3. Suddenly *acting* or *feeling* as if a stressful military experience *were happening again* (as if you were reliving it)?
4. Feeling *very upset* when *something reminded you* of a stressful military experience?
5. Having *physical reactions* (e.g., heart pounding, trouble breathing, sweating) when *something reminded you* of a stressful military experience?
6. Avoiding *thinking about* or *talking about* a stressful military experience or avoiding *having feelings* related to it?
7. Avoiding *activities* or *situations* because *they reminded you* of a stressful military experience?
8. Trouble *remembering important parts* of a stressful military experience?
9. Loss of *interest* in activities that you used to enjoy?
10. Feeling *distant* or *cut off* from other people?
11. Feeling *emotionally numb* or being unable to have loving feelings for those close to you?
12. Feeling as if your *future* will somehow be *cut short*?
13. Trouble *falling* or *staying asleep*?
14. Feeling *irritable* or having *angry outbursts?*
15. Having *difficulty concentrating?*
16. Being “*super-alert*” or watchful or on guard?
17. Feeling *jumpy* or easily startled?

### Combat Exposure Scale (CES)

Please circle the number above the answer that best describes your experience (this scale employs varying Likert-type items ranging from 1-5 with differing measures for each item such as number of times, length of time, or percentage of time).

1. Did you ever go on combat patrols or have other dangerous duty?
2. Were you ever under enemy fire?
3. Were you ever surrounded by the enemy?
4. What percentage of the soldiers in your unit were killed (KIA), wounded or missing in action (MIA)?
5. How often did you fire rounds at the enemy?
6. How often did you see someone hit by incoming or outgoing rounds?
7. How often were you in danger of being injured or killed (i.e., being pinned down, overrun, ambushed, near miss, etc.)?

